# A neural mechanism underlying predictive visual motion processing in patients with schizophrenia

**DOI:** 10.1101/2022.10.11.22280936

**Authors:** Sebastian Scheliga, Rosalie Schwank, Ruben Scholle, Ute Habel, Thilo Kellermann

## Abstract

Psychotic symptoms may be traced back to sensory sensitivity. Thereby, visual motion (VM) processing particularly has been suggested to be impaired in schizophrenia (SCZ). In healthy brains, VM underlies predictive processing within hierarchically structured systems. However, less is known about predictive VM processing in SCZ. Therefore, we performed fMRI during a VM paradigm with three conditions of varying predictability, i.e., predictable-, random-, and arbitrary motion. The study sample comprised 17 SCZ patients and 23 healthy controls. We calculated general linear model (GLM) analysis to assess group differences in VM processing across motion conditions. Here, we identified significantly lower activity in right temporoparietal junction (TPJ) for SCZ patients. Therefore, right TPJ was set as seed for connectivity analyses. For patients, across conditions we identified increased connections to higher regions, namely medial prefrontal cortex, or paracingulate gyrus. Healthy subjects activated sensory regions as area V5, or superior parietal lobule. Since TPJ operates as hub modulating network shifts, aberrant functional connectivity with higher structures may thus reflect a compensatory mechanism co-occurring to impaired TPJ activity in SCZ. In sum, these altered neural patterns provide a framework for future studies focusing on predictive VM processing to identify potential biomarkers of psychosis.

## 1. Introduction

Schizophrenia (SCZ) is a severe mental disorder typically occurring along with positive symptoms such as delusions, hallucinations, or negative symptoms such as diminished emotional expression (McCutcheon et al., 2020). Additionally, SCZ has been linked to sensory processing deficits, particularly for audition (Javitt, & Sweet, 2015) but also for the visual domain (Kogata, & Iidaka, 2018; Yeap et al., 2008). Thereby, a visual dysfunction seems to be crucially involved in the etiology of SCZ whereby deficits in visual motion (VM) processing may even predate the illness onset (Martínez et al., 2018). Therefore, especially the domain of VM processing (Chen, 2011; Li, 2002), and biological motion (Droop et al., 2005; Okruszek, & Pilecka, 2017; Kim et al., 2005) have been studied intensively. However, the underlying neural mechanism and its role in the pathology of SCZ is not comprehensively understood yet.

Using fMRI, studies on visual processing found reduced neural activity in the motion sensitive brain area V5 (corresponding to the middle temporal area) (Lencer et al., 2005). Also, further examinations provide evidence for a global motion processing deficit in SCZ manifesting by reduced brain activity in area V5 (Chen et al., 2008; Martínez et al., 2018). Considering biological motion, functional abnormalities, i.e., reduced activation, were found in superior temporal sulcus (STS) (Kim et al., 2011) and in the temporoparietal junction (TPJ) for SCZ patients (Hashimoto et al., 2014). Another fMRI study on VM perception found reduced V5 activity in patients while, at the same time, revealing increased brain activity in the prefrontal cortex (PFC) that is involved in cognitive processing (Chen et al., 2008). Therefore, the authors concluded that motion processing deficits in patients with SCZ may be associated with reduced brain activity in sensory-related structures and compensatory increased cognitive processing.

According to the predictive coding model of psychosis (Adams et al., 2013; Sterzer et al., 2018), a compensatory effect of cognition-related prefrontal activation may also reflect an enhanced feedback signaling of higher-levels towards sensory brain regions. A similar result has also been reported in an fMRI study investigating VM processing and delusional beliefs in SCZ. Thereby, patients showed increased functional connectivity (FC) between the orbitofrontal cortex presumably encoding cognitive beliefs, and V5 suggesting increased signaling between hierarchical higher brain regions and lower regions which may reflect how higher-level predictions shape lower-level sensory perception (Schmack et al., 2017). Hence, sensory deficits in patients with SCZ and psychotic symptoms as hallucinations may underlie deficits in predictive coding (Horga et al., 2014).

Predictive coding is based on the assumption that perception underlies a hierarchically structured neural system incorporating bottom-up and top-down connections at each level so that sensory input (bottom-up) will be compared to higher order beliefs (top-down), i.e., predictions (Clark, 2013). Imbalances or mismatches between incoming sensory information and predictions are referred to as prediction error. Prediction error signaling seems to be affected in SCZ (McCleery et al., 2018). This is supported by findings pointing to an attenuated mismatch negativity (MMN) in patients with SCZ (Gil-da-Costa et al., 2013). MMN is an event-related negativity usually measured by EEG 150-200ms after the onset of physically deviant auditory stimuli in identical and repeated sequences (Kawakubo et al., 2007; Näätänen et al., 1978). Here, the predictive coding theory postulates that repeated standard tones lead to increased predictability of auditory input for healthy subjects (Lieder et al., 2013) whereas patients with SCZ, due to extraordinary sensory sensitivity, may perceive unpredictability even for very subtle differences in sensory input when they get confronted with a regular stimulus series of consecutive standard tones. Consequently, the response to a deviant tone contrasted to standard stimuli yields a diminished negativity (MMN) as compared to healthy controls.

It has been demonstrated that unpredictable VM is based on hierarchical processing (Kellermann et al., 2017). Since SCZ is associated with deficits in VM processing (Chen, 2011; Li, 2002; Martínez et al., 2018) it is possible that these impairments underlie an imbalance between connections from different hierarchical processing levels (Adams et al., 2013). Sensory MMN deficits are a robust pathological feature in SCZ (Umbricht, & Krljes, 2005), particularly for the visual domain (Kremláček et al., 2016; Neuhaus et al., 2013; Urban et al., 2008). Accordingly, an attenuated MMN to moving visual stimuli may be reflected by altered activity in brain regions as area V5 (Lencer et al., 2005), STS (Kim et al., 2011), and TPJ (Hashimoto et al., 2014).

Additionally, activation deficits may occur along with an aberrant FC. In SCZ, altered neural connections are suggested to represent a key feature of the disorder (Li et al., 2017) that even occurs in patients at an early-stage (Hummer et al., 2020). To date, some fMRI studies have investigated predictive signaling in SCZ (e.g., Schmack et al., 2017). However, evidence on neural alterations during predictive VM processing in particular is rather scarce. Therefore, in our fMRI study we use a VM paradigm with decreasing predictability to assess aberrant sensory processing in SCZ.

The present study aims at providing new knowledge about the neural correlates of VM processing in SCZ. Our examination is based on the conceptual idea that psychosis underlies compromised predictive signaling and consequently suboptimal prediction errors (Adams et al., 2013). Therefore, using fMRI we assessed brain activation and FC during a visual paradigm that comprises three different motion conditions with constantly decreasing predictability. Since prediction error in SCZ may be increased independently of the degree of predictability (Lieder et al., 2013; McCleery et al., 2018), we expect that the predictability of sensory (visual) stimuli has less impact on SCZ patients compared to healthy controls. Therefore, we assume that the predictability of the visual stimulus is reduced in patients manifesting in smaller activation differences between the three motion conditions. We expect these activation deficits to occur in brain regions that have been associated with VM processing before, i.e., area V5 (Martínez, et al., 2018), STS and TPJ (Patel et al., 2021).

Since especially TPJ has been linked to the modulation of attention (Corbetta et al., 2000; DiQuattro et al., 2014), we expect that particularly patients with SCZ, relatable to attentional deficits (Adams et al., 2013; Matsumoto et al., 2018), may have altered activity in this brain region. Finally, for TPJ we expect impaired functional connections to other brain regions involved in VM processing while an abnormal prediction error processing may rather facilitate increased functional coupling with higher brain regions such as prefrontal cortex (PFC) (Yaple et al., 2021). Referring to a theoretical framework based on the predictive coding theory in psychosis, our fMRI study investigates the neural correlates of predictive VM processing in patients with SCZ.

## 2. Methods

### 2.1 Participants

Our study sample comprised 23 healthy control subjects and 17 patients who met the International Classification of Disease (ICD-10) (World Health Organization, 1992) criteria for a diagnosis from the SCZ spectrum. There were ten patients with a SCZ diagnosis (F20.0), one with schizoaffective disorder (F25.0), two patients with undifferentiated SCZ (F20.3), one with SCZ simplex (F20.6), one with psychosis induced through psychoactive substances (19.5), and one patient with a diagnosis of residual SCZ (F20.5). Using the structured clinical interview for axis I disorders (SCID) (First, & Gibbon, 2004) for the control group, none of the subjects had a psychiatric or neurological disorder or received psychotropic medication. A medication list comprising the type and doses of antipsychotics taken by the patients can be found in the supplementary material (Table S1). For all participants, demographics and clinical data can be found in table 1. The Positive and Negative Syndrome Scale (PANSS) was used to assess SCZ psychopathology in patients. Notably, concerning sex and age the groups did not differ significantly (Table 1).

**Table 1.** Participant demographic and clinical data.

|  | Healthy Controls | SCZ Patients | *p*-value |
| --- | --- | --- | --- |
| N (m/f) | 23(16/7) | 17(11/6) | - |
| Age<sup>a</sup> | 36.22 (11.65) | 37.35 (11.77) | 0.763 |
| Duration of illness (years)<sup>a</sup> | - | 8.42 (8.16) | - |
| PANSS positive symptoms<sup>a</sup> | - | 13.59 (5.19) | - |
| PANSS negative symptoms<sup>a</sup> | - | 14.41 (5.75) | - |
| PANSS general symptoms<sup>a</sup> | - | 28.00 (6.41) | - |
| PANSS total score<sup>a</sup> | - | 54.71 (12.51) | - |

**Abbreviations.** PANSS, Positive and Negative Syndrome Scale
**Notes.** <sup>a</sup> Mean (Standard deviation)

### 2.2 Stimuli and task

We presented visual stimuli using an MR-compatible googles system (Resonance Technology Company Inc., Los Angeles, USA). Within the visual field of subjects, the screen covered 25° x 19° with a resolution of 800 × 600 pixels. We used the Presentation software (Neurobehavioral Systems Inc., Berkely, USA) to control the application of stimuli. Visual stimuli comprised a black frame (24° x 16°) on a white background with a filled circle in the middle (or ‘ball’). The black ball was presented stationary within the black frame during the baseline condition. Both conditions (‘count’ and ‘active’) started with the ball’s position at the center of the screen. For the following blocks (again for both conditions) the ball’s initial position was the same as its last position from the previous baseline condition. In total, we conducted 30 experimental blocks. In each block, while not leaving the frame the ball moved constantly with a speed of 6° per second for 20, 20.5 or 21.5 seconds. We inserted a baseline with a mean length of 10 seconds between two subsequent experimental blocks. At baseline, the ball stopped moving and stayed at the last position. After a jitter of 0, 0.5 or 1.5 seconds, the ball began to move from its last position.

Each run comprised 30 experimental blocks and was divided into three different conditions. For each run, we used a pseudorandomized order as well as counterbalanced durations and jitters. In the predictable condition, the ball changed its motion direction only if it touched the frame’s border where the angle of dip corresponded to the emergent angle. During the random condition, the ball’s rebound varied randomly and did not correspond to the incident angle with the constraint that the ball stayed within the frame making this condition less predictable than the aforementioned. The least predictable was the arbitrary condition. Within this condition, changes of motion direction not only occurred with contacts of the ball with the cushion but also in random intervals in the middle of the frame. Taken together, predictability of motion thus decreased from the predictable over the random to the arbitrary condition (Figure 1).

**Figure 1.**
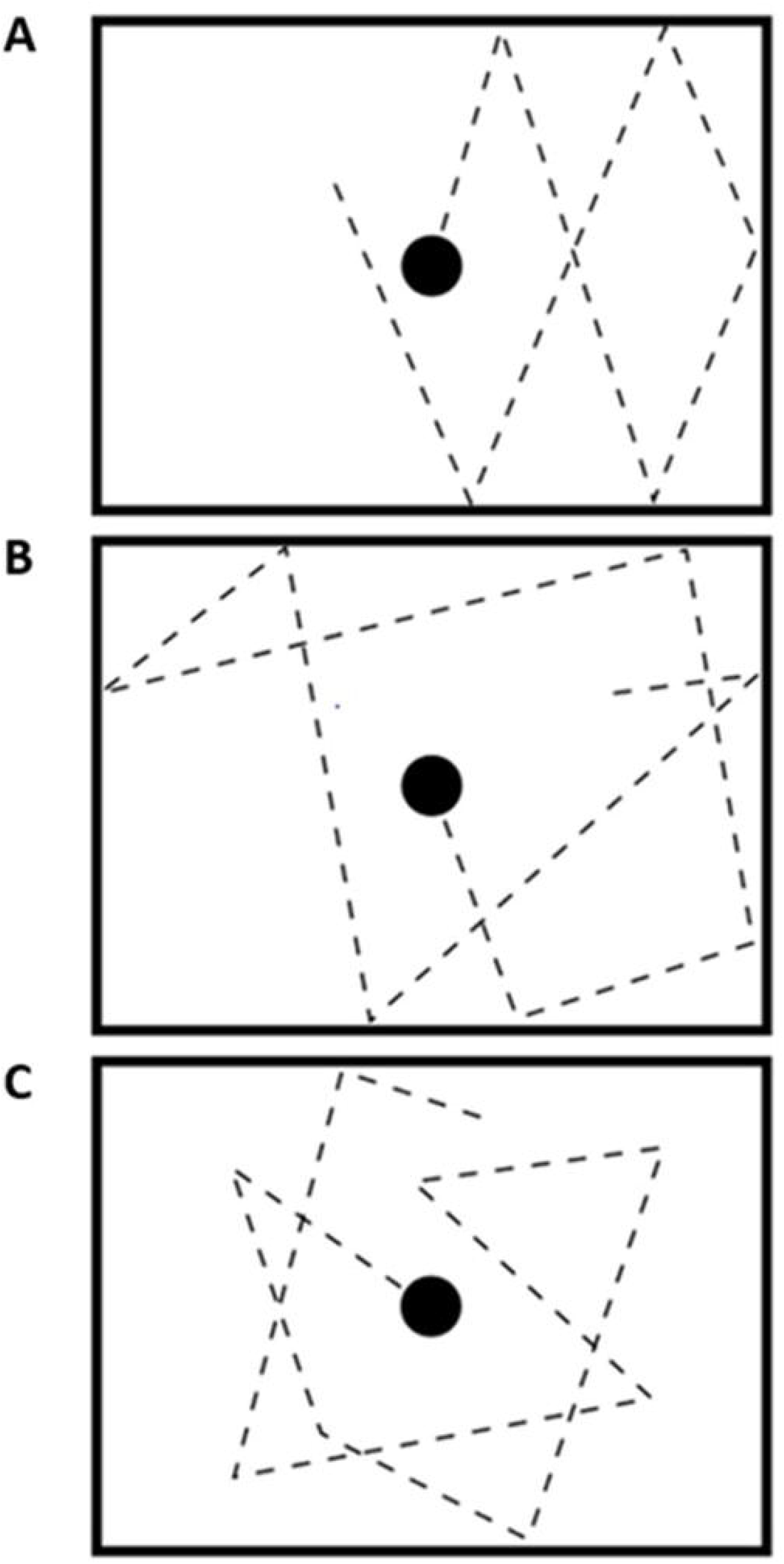

In total, there were two different runs. Thereby, in each run the participants were instructed to keep track of the ball and to attend to its changes in motion direction. Only considering the response mode, the runs differed. In the active run, the subjects had to press a button with the right index finger when they perceived a change in the motion direction. Whereas, in the passive run they had to keep track of the ball and (silently) count the number of motion direction changes only. After each trial subjects had to choose the correct answer out of four alternatives after each block. During the count run, participants had 5 s to give an answer before the next block started. Notably, the whole procedure was adapted from our former study on VM processing (see also Kellermann et al., 2017).

### 2.3 Data acquisition

We performed functional magnetic resonance imaging (fMRI) using a Siemens 3T MRI scanner. For the active-condition (no response), per subject we acquired 520 functional images using a T2*-weighted echo-planar imaging (EPI) sequence that covers the whole brain with 33 axial slices each having a thickness of 3.4 mm (a gap of 0.51 mm between the slices). For the count-condition (response) we acquired 609 EPI images using the same parameters as in the active-condition. The resolution for each slice was 64 × 64 pixels with a field view of 200 × 200 mm^2^ resulting in a voxel size of 3.125 × 3.125 × 3.4 mm^3^. Since the echo-time (TE) was 30 ms, and the flip angle amounted to 75° while the repetition time (TR) was 1800 ms, we thus had an acquisition time of 16 min for the active-condition and an acquisition time of 18 min and 27 s for the count-condition. Due to T1 stabilization effects, we discarded the first three images for each run. After the functional measure, we also acquired an anatomical image with T1-weighted magnetization prepared rapid gradient echo (MPRAGE) sequence with a resolution of 1 × 1 × 1 mm^3^ (TR: 1900 ms, TE: 2.52 ms, flip-angle: 9°).

### 2.4 Data preprocessing

We preprocessed fMRI data using the Functional Connectivity toolbox (CONN) (https://www.nitrc.org/projects/conn, version 19.c) implemented in SPM12. Here, we conducted the default preprocessing pipeline, i.e., fMRI data were realigned and unwarped, followed by functional slice time correction, structural and functional normalization, and segmentation into gray matter, white matter, and CSF tissue. We identified potential outlier scans from the observed global BOLD signal and the amount of subject motion in the scanner. Thereby, scans were marked as potential outliers if framewise displacement was above 3mm or global BOLD signal changes above 5 standard deviations. Finally, functional images were smoothed using a spatial convolution with an 8 mm full width half maximum (FWHM) Gaussian kernel. We further used CONN’s default denoising pipeline combining linear regression of potential confounding effects in the BOLD signal, and temporal band-pass filtering in order to remove any confounding influence on the signal. Functional data were resampled and interpolated to a spatial resolution of 2×2×2 mm^2^ voxels. We focused on noise components from white matter, CSF, estimated subject motion parameters, scrubbing, as well as session- and task effects (e.g., Emam et al., 2019). To focus on slow frequency fluctuations, the functional data were band-pass filtered afterwards (0.008 – 0.09 Hz) to minimize the influence of noise sources.

### 2.5 General linear model (GLM) analyses

After preprocessing of fMRI raw data, we modeled the two runs per subject by convolving the boxcar functions of the three motion conditions per run with the canonical hemodynamic response. Thereby, we modeled the phase when the ball was presented stationary as implicit low-level baseline. On the first level, the design matrix thus resulted in six predictors (2 runs by 3 conditions). We used these predictors as regressors in the GLM. Here, the realignment parameters and intercepts were treated as covariates of no interest. Accounting for low-frequency drifts, we used a high-pass filter with a cut of 128 seconds. In addition, by removing estimated first-order autoregressive effects of the time-series, temporal autocorrelations were considered as well. For the second levels statistics, we used the contrast estimate images of each subject and each condition and calculated a three-way repeated measures ANOVA. This model comprised the within-subject factor *predictability* with three levels (predictable, random, arbitrary), the within-subject factor *task* with two levels (active and passive), and the between-subject factor group with two levels (healthy controls, SCZ patients). Notably, for the active and the passive level we do not expect significant differences considering brain activation during VM processing (Kellermann et al., 2017). Therefore, this factor was omitted for the analyses. We set the threshold for rejecting the null hypothesis to p < 0.05 (FWE) at the voxel level for multiple comparisons per contrast (extent threshold of 100 continuous voxel).

### 2.6 Seed-based connectivity analysis

We used a seed-based correlation approach to compare FC during VM processing between SCZ patients and healthy controls. Based on our finding from the GLM analysis (see results section), we placed the seed for FC analysis within the right TPJ [50, -40, 18]. Using the CONN toolbox, a seed to voxel analysis was performed comparing the temporal correlation of the BOLD signal to all remaining voxels in the brain. Thereby, for each analysis we set the right TPJ’s MNI coordinates as seed region with a spherical radius of 6 mm. Fisher transformed z scores were used considering multiple comparisons. In total, we conducted one connectivity analysis for each of the three VM conditions (predictable, random, and arbitrary). Using a GLM, the analysis was conducted across all study subjects to identify significant TPJ-connections at the first level (Single subject level). After the first level analysis, the results were further used for second level analysis aiming to identify significant group differences in FC between SCZ patients and healthy controls. For second level analysis, an unpaired t-test was calculated with a voxel wise cluster-forming threshold set at *p* < 0.001 uncorrected in combination with an extent threshold corresponding to *p* < 0.05 family wise error (FWE) corrected at the cluster level. For each motion condition, we defined the contrast weights 1 for healthy controls and –1 for SCZ patients. Thus, positive t-values indicate increased functional connections in control subjects while negative values points to higher FC in the patient group.

## 3. Results

### 3.1 GLM

Across all conditions, we tested the main effect for the factor group comparing SCZ patients with healthy controls. In patients, we found significantly lower brain activity (*F* = 23.05, *p* < 0.05 FWE) within two clusters in the left TPJ [-52, -52, 16] (51 voxels), and in the right TPJ [50, -40, 18] (220 voxels). Notably, the finding on the right TPJ also survived a more conservative threshold correction of *p* < 0.001 (FWE) (Figure 2). The interaction effect group x predictability was not significant for the FWE corrected threshold at *p* < 0.05 (*F* = 13.64). However, using a more liberal threshold, activation in the right TPJ yielded statistical significance (*F* = 7.12, p < 0.001 uncorrected).

**Figure 2.**
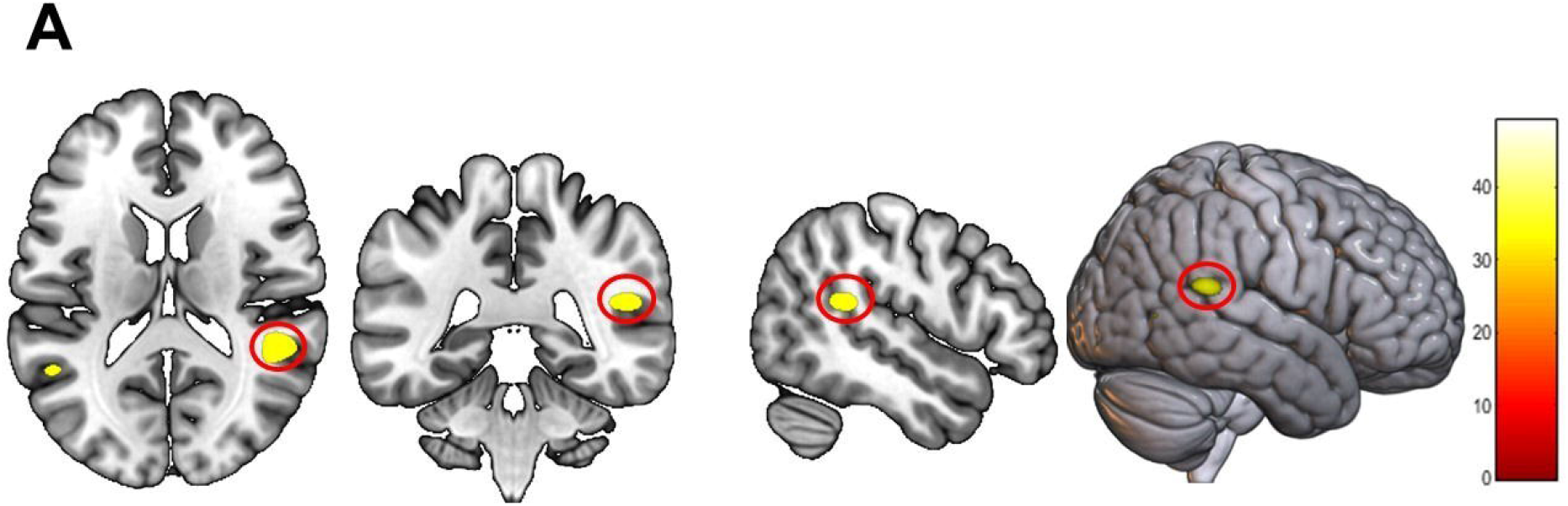
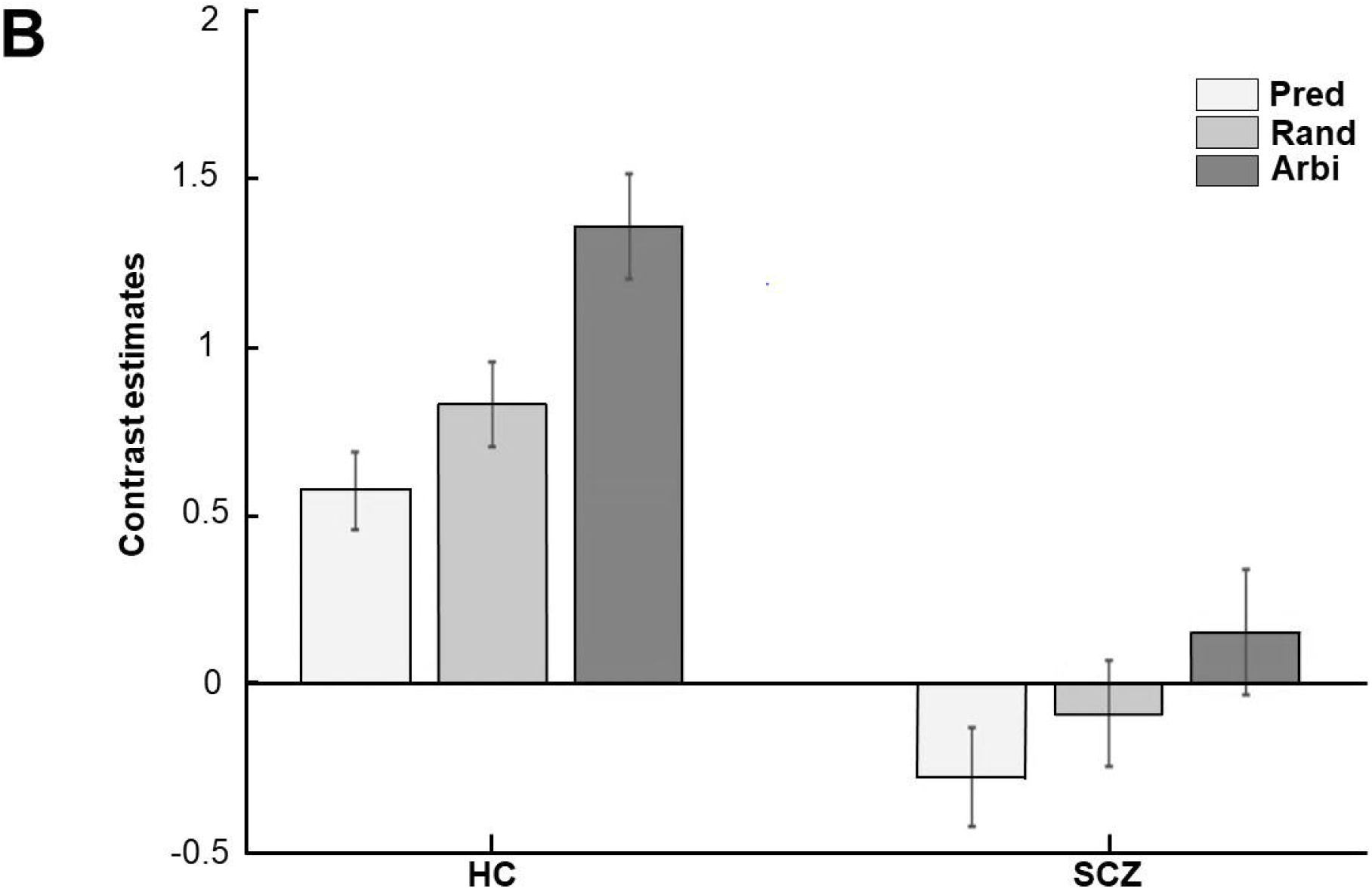

### 3.2 Seed to voxel connectivity

In the **predictable** condition, for patients we identified higher connectivity with the TPJ within the precuneus, left SOC, and mPFC while healthy controls showed higher connectivity in right V5 (see Table 2 and Figure 3).

**Table 2.** Clusters of between-group seed connectivity differences for **predictable**.

| Region | k | Peak MNI coordinates |  |  | Peak T score |
| --- | --- | --- | --- | --- | --- |
|  |  | x | y | z |  |
| R V5 | 337 | 54 | -56 | -4 | 5.54 |
| R precuneus | 263 | 6 | -58 | 24 | -4.41 |
| L SOC | 216 | -58 | -68 | 20 | -4.35 |
| L mPFC | 131 | -4 | 44 | -18 | -3.97 |

**Abbreviations.** R, right; L, left; mPFC, frontal medial cortex; V5, lateral occipital cortex, SOC, superior occipital cortex

**Figure 3.**
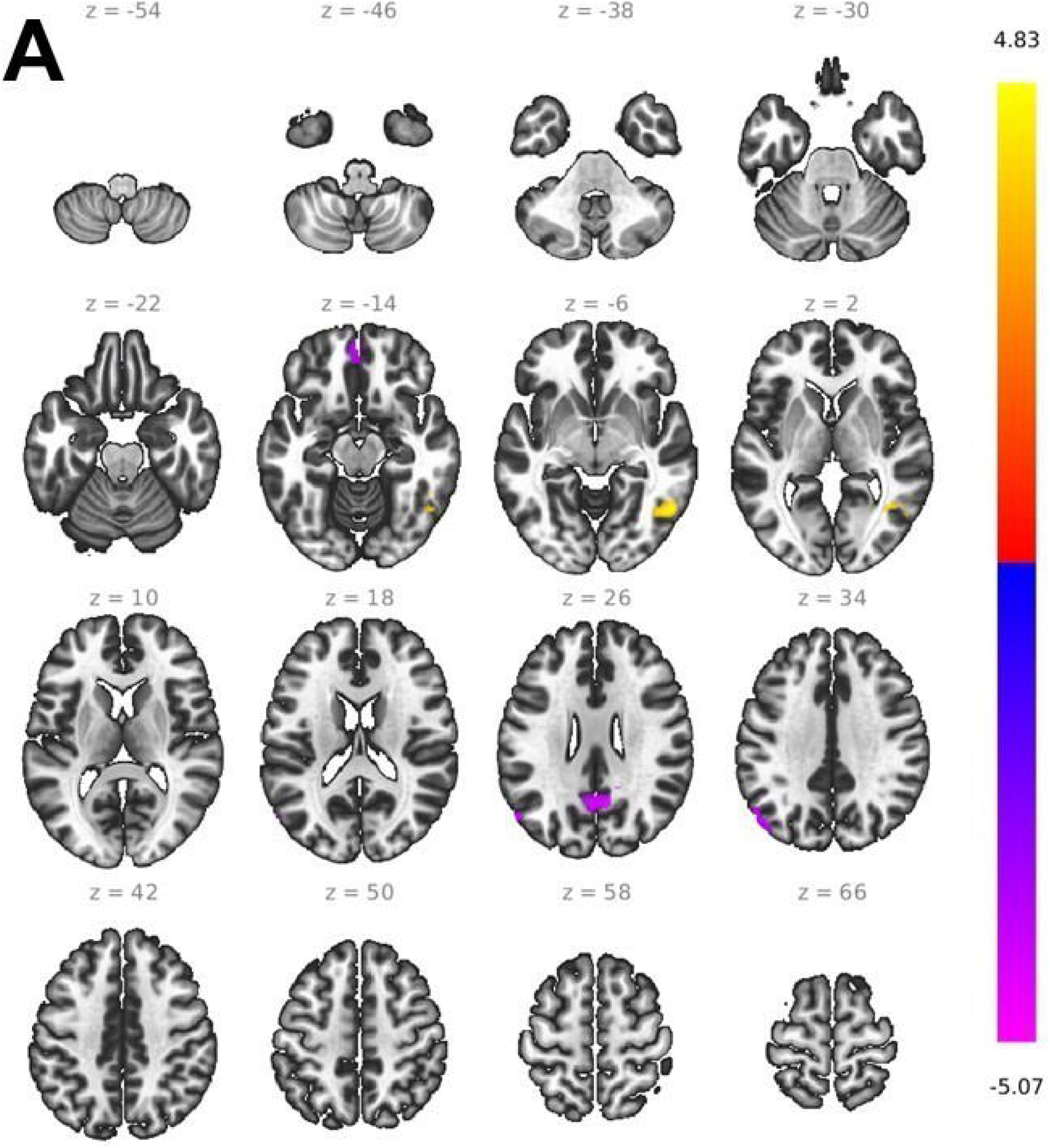
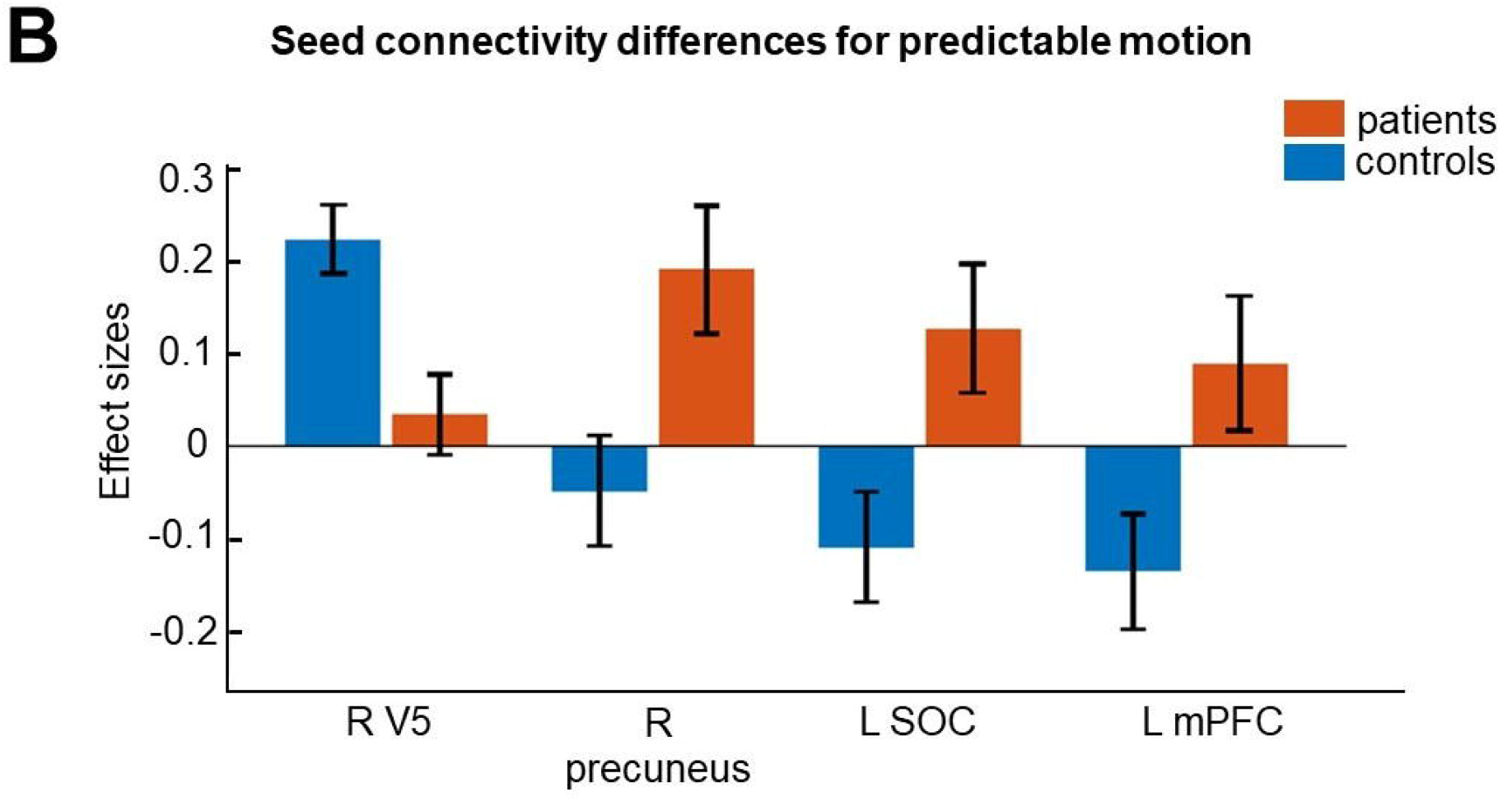

In the **random** condition, patients showed higher connectivity with the TPJ in the mPFC, left frontal pole, and right paracingulate gyrus (PcG) while TPJ-based connectivity for healthy controls was stronger with the right SPL, right ITG, and the right SFG (see Table 3 and Figure 4).

**Table 3.**
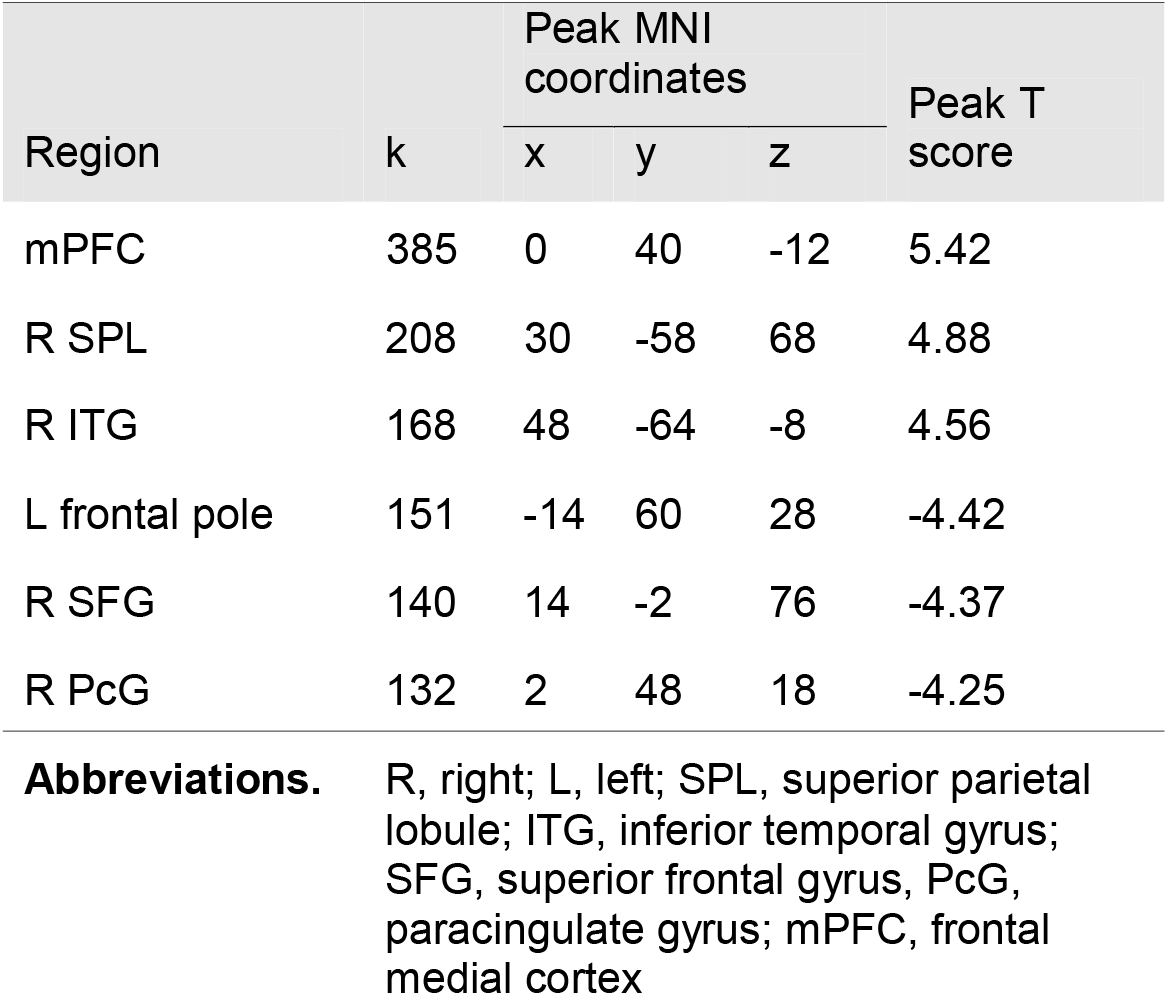
Clusters of between-group seed connectivity differences for **random**.

**Figure 4.**
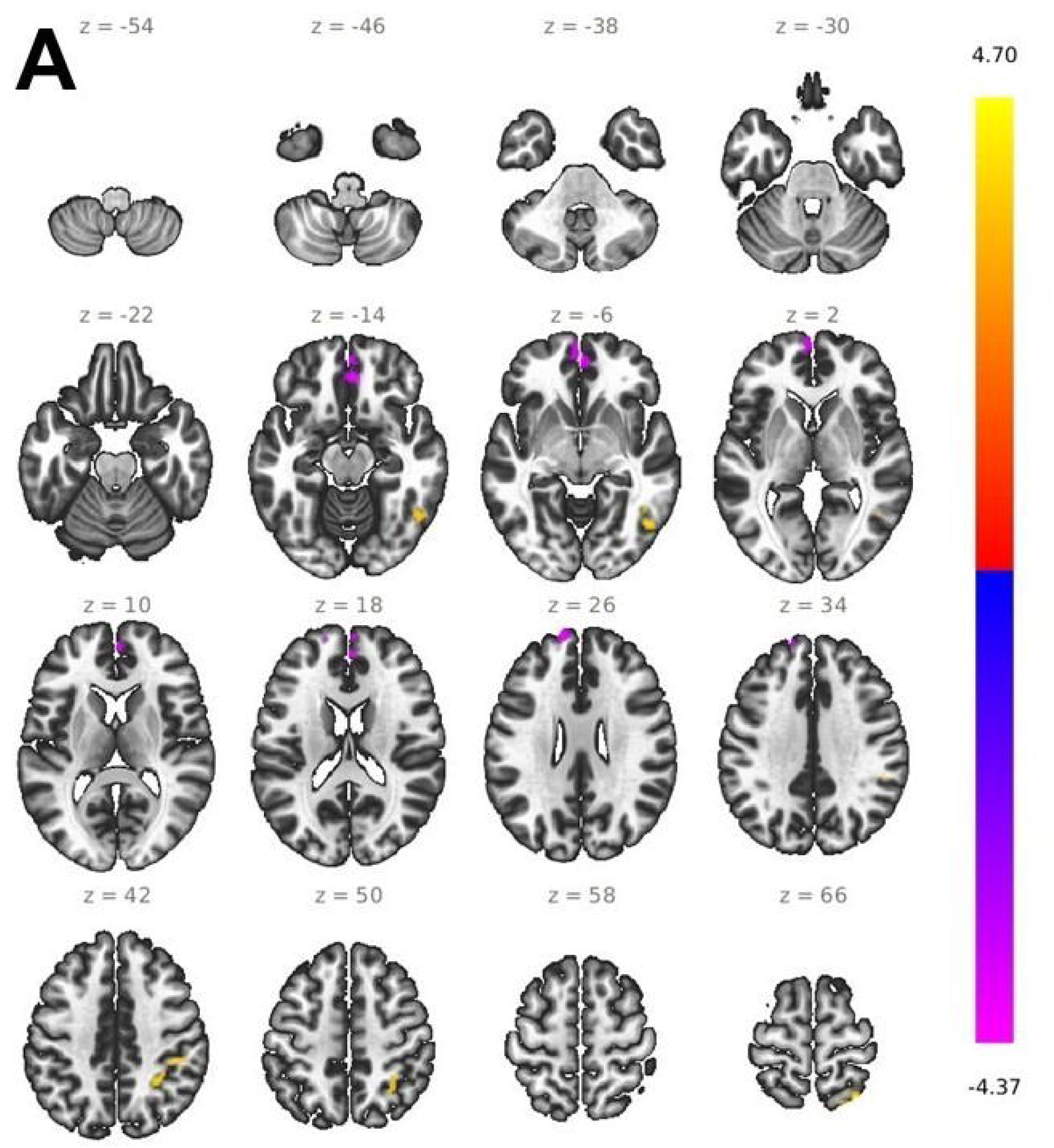
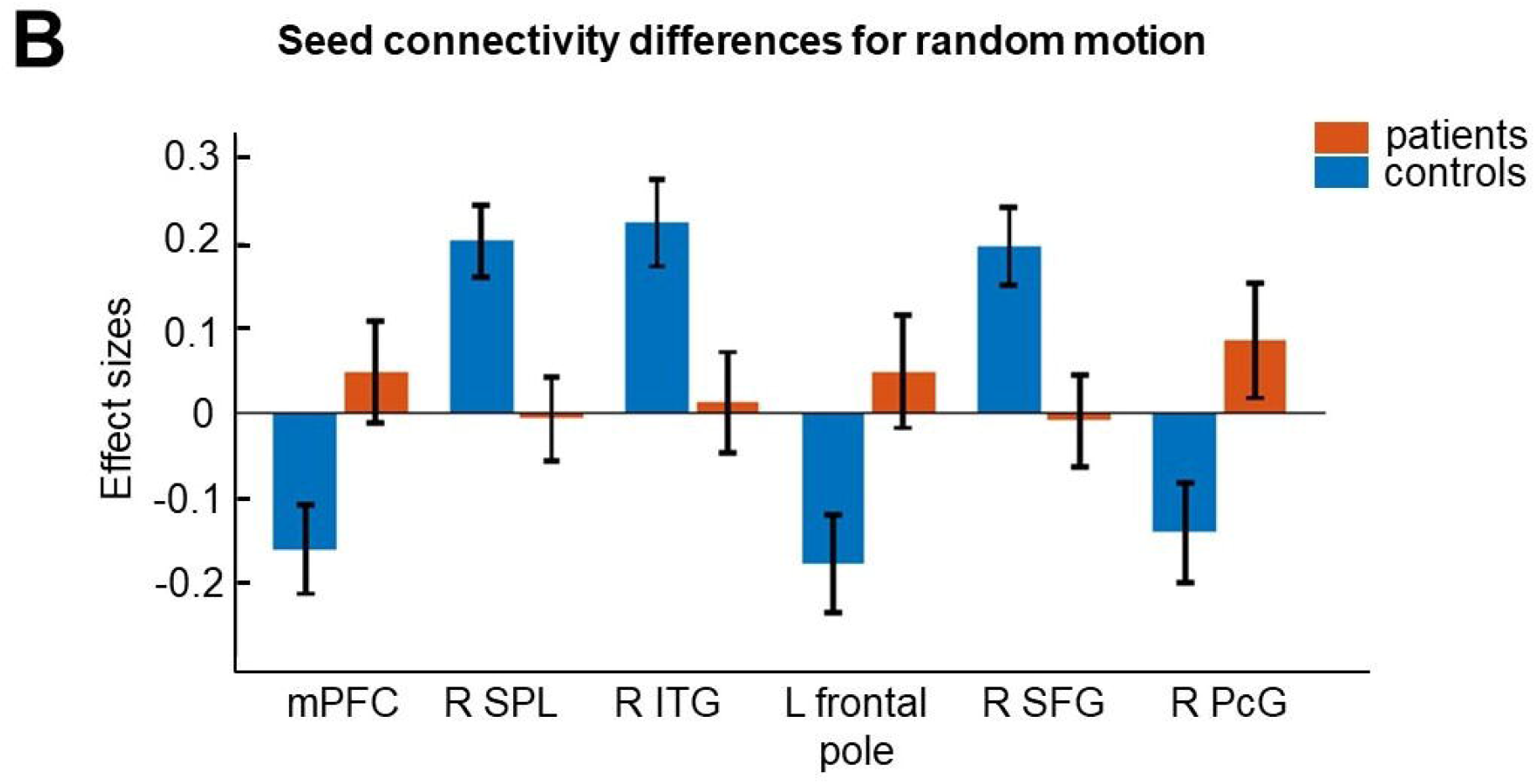

In the **arbitrary** condition, for each brain region we found higher connectivity for the TPJ in SCZ patients (see Table 4 and Figure 5).

**Table 4.** Clusters of between-group seed connectivity differences for **arbitrary**.

| Region | k | Peak MNI coordinates |  |  | Peak T score |
| --- | --- | --- | --- | --- | --- |
|  |  | x | y | z |  |
| R cerebellum | 481 | 28 | -72 | -38 | -6.82 |
| L SFG | 285 | -2 | 34 | 60 | -5.77 |
| L PCG | 222 | -14 | 40 | 10 | -5.02 |

**Abbreviations.** R, right; L, left; SFG, superior frontal gyrus; PcG, paracingulate gyrus

**Figure 5.**
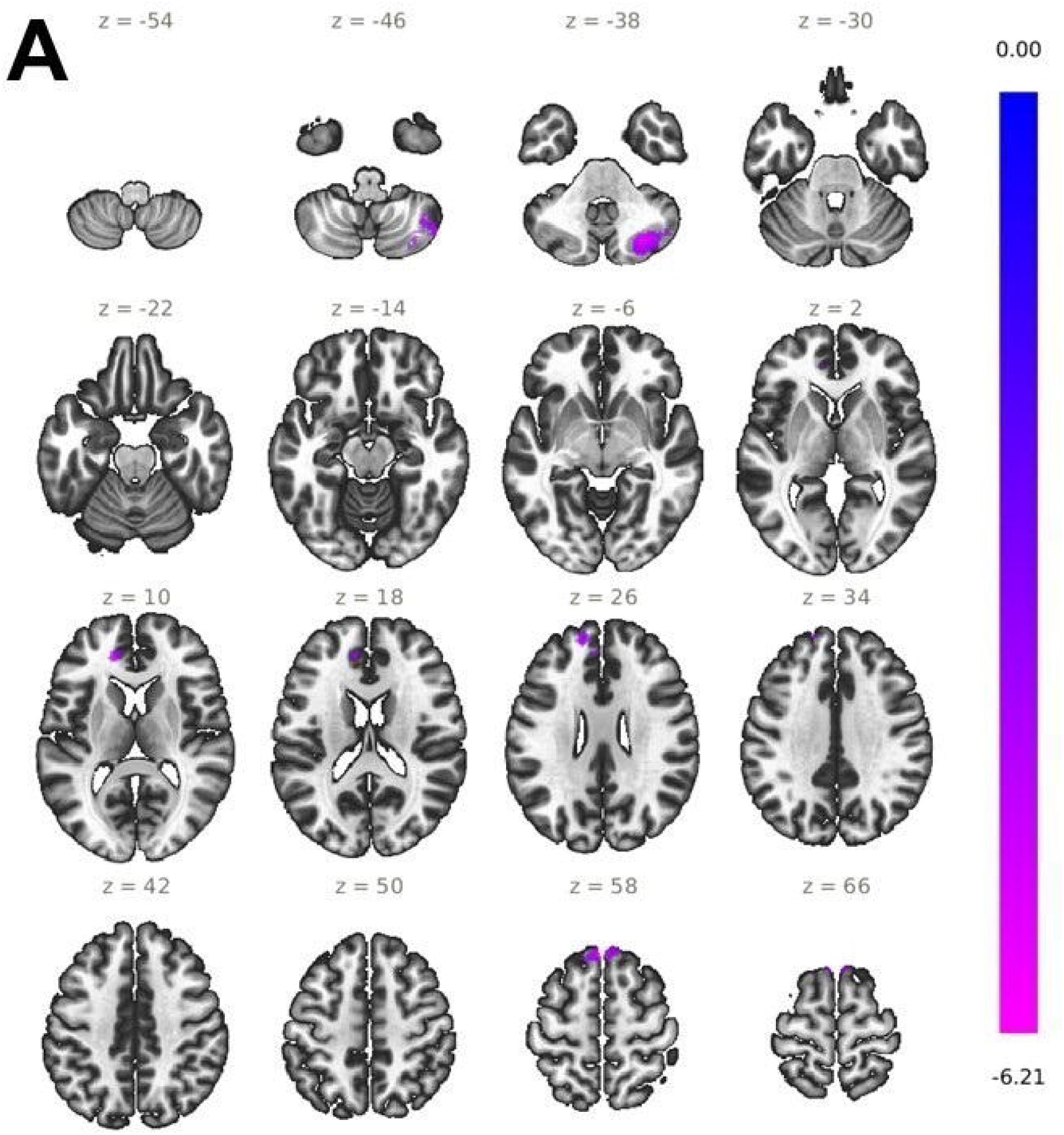
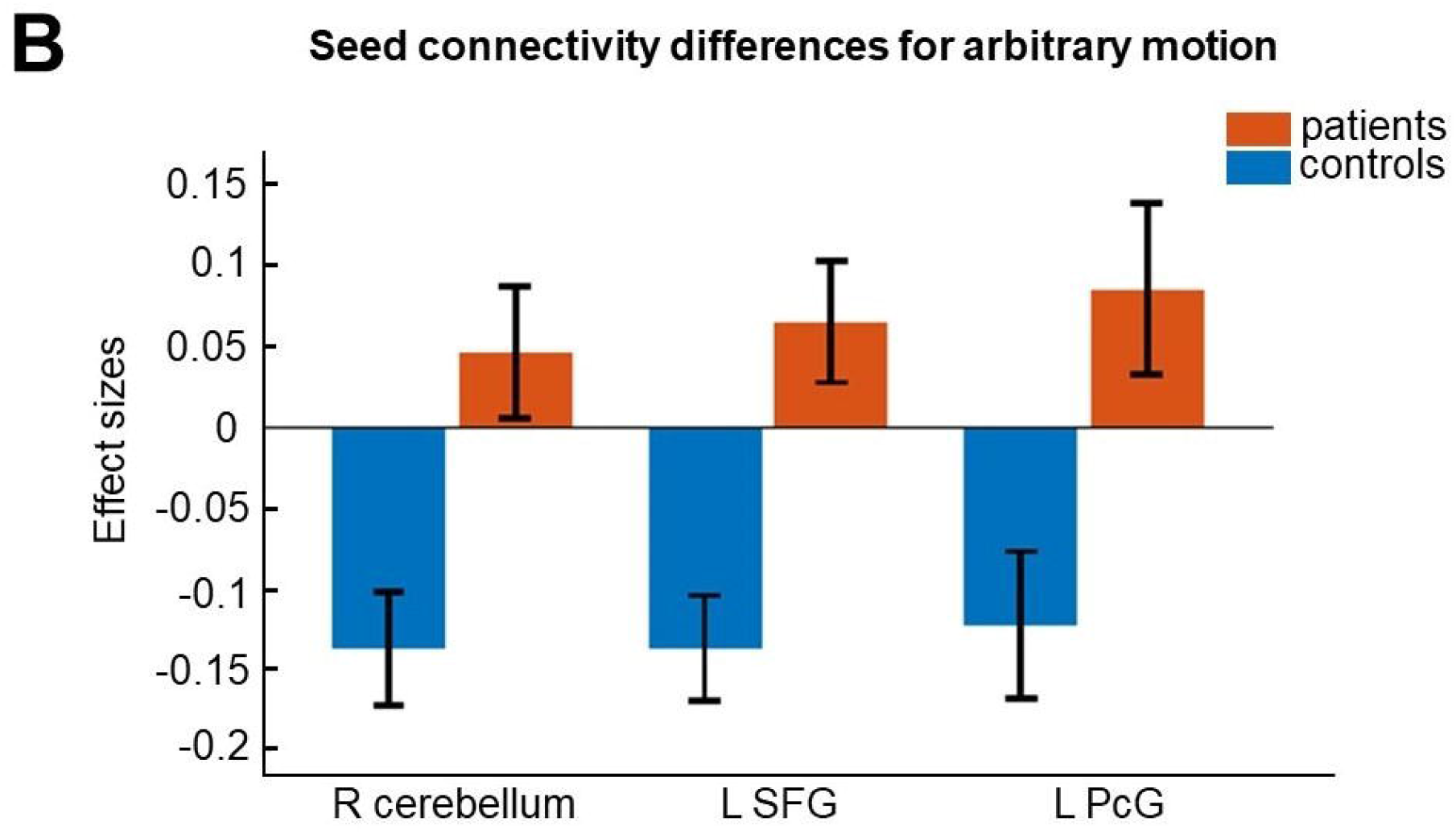

### 3.3 Psychopathology correlations

We performed a correlation analysis to account for a link between results at the neural level and psychopathology scores within the SCZ group. Therefore, we considered the patients’ PANSS scores (positive symptom scale, negative symptom scale, general symptom scale, and total score) and correlated them with the beta-values of the right TPJ [50, -40, 18] that we received from the GLM analysis. For each correlation a significance threshold of *p* < 0.001 (uncorrected) was used. We could not find significant TPJ correlations with the positive symptom scale (*r* = 0.2712; *p* = 0.2923), negative symptom scale (*r* = 0.2651; *p* = 0.3037), general symptom scale (*r* = 0.2223; *p* = 0.3912) and with the total score (*r* = 0.3198; *p* = 0.2108).

## 4. Discussion

The purpose of the present fMRI study was to investigate neural mechanisms underlying VM processing in patients with SCZ. Therefore, we conducted a VM paradigm with three different conditions comprising predictable-, random-, and arbitrary motion, respectively. As expected, across all motion conditions we found significantly reduced brain activation in right TPJ for SCZ patients compared to healthy controls. Using right TPJ as seed region, we further conducted FC analyses for each motion condition. For predictable motion, we found higher functional connections to right precuneus, and left mPFC in SCZ patients. In the random motion condition, we identified higher connectivity to the left frontal pole, right SFG, and right PcG in the patient group. Interestingly, observing arbitrary motion, the least predictable condition, patients showed increased FC to the right cerebellum, left SFG, and left PcG whereas there was no brain region showing higher functional coupling in healthy subjects. Taken together, these findings reflect a neural pattern of aberrant functional TPJ connectivity during predictive VM processing that may be a characteristic feature in SCZ.

Across all motion conditions, we identified significantly lower TPJ activity in SCZ patients. This finding is in line with previous studies on SCZ linking dysfunctional TPJ activity to deficits in attentional modulation (Matsumoto et al., 2018). Traditionally, the TPJ is involved in stimulus-driven reorienting of attention (Corbetta et al., 2000) while especially the anterior portion of this brain region may be involved in target detection (Patel et al., 2019). However, the TPJ may also exert a contextual updating function instead of initiating bottom-up attentional reorienting only (DiQuattro et al., 2014; Geng, & Vossel, 2013). Interestingly, we did not identify significant group differences for the STS. The absence of STS activity might be due to its involvement in higher order biological motion rather than in more general VM processing (Matsumoto et al., 2018).

In each motion condition, TPJ activity was higher for healthy controls supporting a previous study which has shown that increasing uncertainty, i.e., lower movement predictability enhances the neural activity within the right TPJ, suggesting a crucial role for this region in (visual) predictive coding (Jakobs et al., 2009). Referring to the ‘Bayesian brain hypothesis’, predictive coding is the continuous process of updating prior beliefs about the state of the environment with actual sensory input. In this context, attention, can enhance the precision of sensory input which is the same as reducing the uncertainty of this input (Clark, 2013; Friston, 2010; Knill, & Pouget, 2004). In our study, continuously increasing TPJ activity from predictable-, through random-, to arbitrary motion suggests that decreasing predictability requires steadily updating of prior beliefs, a mechanism of predictive coding that may manifest by enhanced neural activity in the TPJ (Jakobs et al., 2009). On the other hand, across all motion conditions the significantly lower TPJ activity in the patient group may suggest a higher prediction error. Basically, unexpected stimuli are associated with a larger prediction error that should result in enhanced connectivity from lower to higher brain regions in the processing hierarchy (Adams et al., 2013). In a previous fMRI study, using the same VM paradigm as in the present study, we tested modulatory effects of hierarchical predictive coding in the visual system by calculating dynamic causal modelling (DCM) (Kellermann et al., 2017). Here, we identified enhanced strength of forward connectivity from V1 to V5 supporting the hypothesis of augmented bottom-up processing of prediction error with decreasing motion predictability (Kellermann et al., 2017).

In the present study, we identified significantly increased TPJ connectivity with the lateral occipital cortex, referred to as V5 (Peigneux et al., 2000) for healthy controls during predictable motion. This FC was significantly lower in the patient group providing further evidence for an imbalance in the precision of sensory processing, presumably yielding a diminished ability to predict VM. Comparably to the MMN where repeated tones lead to high predictability of auditory stimuli (Gil-da-Costa et al., 2013; Tada et al., 2019), excessive sensory sensitivity in SCZ patients presumably lead to a prediction error even for predictable stimuli, resulting in a diminished MMN-response (Winterer, & McCarley, 2011). Although, with our connectivity approach, we cannot make assertions about the direction of influence, in the control group, however, we found higher TPJ coupling with lower-level brain regions as V5 that may indicate an enhancement of bottom-up connectivity during VM for healthy subjects.

In healthy brain functioning, right TPJ may be a hub region for coordinating predictive processing in the human brain that is activated during the prediction of environmental stimuli. SCZ patients, however, may have a deficit in activating TPJ, while at the same time compensatory increased TPJ connections with hierarchical higher brain regions (e.g. mPFC, SFG, PCG) may occur that may also reflect a prediction error. This is supported by the assumption that patients with SCZ have a hyper-sensitivity to sensory input that also underlies attentional processes (Adams et al., 2013). Notably, attentional shifting processes are known to be modulated by anterior right TPJ (Krall et al., 2014). Moreover, this brain structure represents a key region supporting contextual updating that is indispensable in attentional processes. Here, TPJ signals are assumed to reflect (slow) post-perceptual processing and adjustment of top-down expectations (DiQuattro et al., 2014). Thus, a dysfunction of this brain region may lead to aberrant FC with higher brain regions reflecting a large prediction error during VM.

In addition to that, across motion conditions, healthy controls showed higher functional TPJ coupling with area V5, SPL, and ITG, regions activated in attentive tracking of moving objects (Culham et al., 1998) This mechanism is impaired in patients with SCZ (Chen, 2011; Lencer et al., 2005). On the other hand, in patients we identified increased FC with brain regions also related to mentalizing and control processes (e.g., mPFC) (Green et al., 2015). Notably, a similar synchronization pattern has also been reported in a previous fMRI study suggesting that reduced TPJ activity may be either due to a failure in the bottom-up flow of visual information or due to the decrease of signal processing as a consequence of increased top-down input from frontal areas (Patel et al., 2021). The authors suppose healthy brain functioning may require negative coupling between the TPJ and higher structures. A phenomenon that has been referred to as decreased network anticorrelation, which has been reported to be reduced in SCZ (Baker et al., 2014). Therefore, it is possible that our connectivity results may reflect the consequence of a decreased functional anticorrelation between the TPJ and frontal brain regions in SCZ.

Abnormal connectivity of brain networks is assumed to be involved in SCZ (Andreasen et al., 1998; Andreasen et al., 1999; Stephan, Friston, & Frith, 2009; Friston, 1998) while two intrinsic anticorrelated networks are of special interest for the pathophysiology of the disorder, the task-negative network (TNN) also referred to as default-mode network, and the attentional task-positive network (TPN) (Broyd et al., 2009; Liu et al., 2012; Williamson, 2007; Zhou et al., 2007). Both, TPN and TNN, are two reciprocally competitive brain networks, where TPN activation requires TNN deactivation and vice versa (Buckner et al., 2008; Fox et al., 2005; Liu et al., 2012). In SCZ, studies have reported brain network abnormalities in TNN with a lack of suppression resulting in a hyper-activation of this network that may contribute to cognitive impairments in SCZ (Anticevic et al., 2012; Ramkiran et al., 2019). During our VM task, in patients we identified increased TPJ coupling with TNN regions (e.g., precuneus, mPFC, SFG) that may suggest aberrant FC between default-mode brain structures in SCZ (Wolf et al., 2009). Notably, one crucial function of anterior TPJ is modulating activation changes between the task-negative default mode- and the task-positive salience network (Patel et al., 2019). The coordination between these brain networks can be seen as an integration process between internal information and external influences that is impaired in SCZ.

Besides that, SCZ patients showed increased TPJ connectivity to theory of mind (ToM)-regions like PcG or cerebellum (Carrington, & Bailey, 2009; Gallagher, & Frith, 2003; Van Overvalle et al., 2014). In SCZ, studies suggest that patients may have a ‘hyper-ToM’ attitude to overattribute not only intentions to persons but also to objects (Abu-Akel, & Bailey, 2000; Blakemore et al., 2003). Here, SCZ patients may have a hyperactive intention detector that is activated also by physical events leading patients to attribute intentions and goals to physical objects (Bara et al., 2011; Ciaramidaro et al., 2014; Walter et al., 2009; Frith, 2004). This hyper-intentional mode of cognition, treating objects as persons, is reflected in increased brain connectivity in SCZ (Whitfield-Gabrieli et al., 2009). For our VM paradigm, it may be possible that patients perceive the balls’ motion as intentional. Thus, enhanced animacy perception of a physical object may be a further explanation for the increased TPJ connectivity to social brain structures in the patient group (Santos et al., 2010). However, since we did not collect behavioral data related to the participants’ perceived animacy of the balls’ movement we were not able to directly assess this interpretation. Therefore, future studies could also include animacy related questionnaires.

Finally, we calculated correlation analyses to investigate the relationship between our results on the neural level and psychopathology scores. However, we did not find significant correlations between TPJ activity and symptomatology measured by the PANSS. This is in line with previous studies investigating TPJ activations in SCZ patients. Here, the authors could not detect significant correlations between the (right) TPJ and each PANSS score (Salgado-Pineda et al., 2022; Vercammen et al., 2010). One possible explanation is that we failed to identify significant correlations because the distribution of PANSS total score was very heterogeneous within the patient group (range from 36 to 83). This in turn might be traced back to the wide distribution of illness duration (range from 1 to 27 years). However, according to our VM paradigm, we assume that reduced TPJ activity we found in our study may be rather related to a general sensory processing deficit than to the patients’ symptoms.

### 4.1 Limitations

There are several limitations of our study that should be considered for future investigations. Compared to other studies with larger sample sizes, it must be mentioned that our clinical sample consisted of 17 SCZ patients only. Thereby, most patients took neuroleptics at the time of the measurement so that an effect of antipsychotic medication cannot be ruled out. Here, unmedicated patients would be desirable. Also, the patient sample comprised different diagnosis from the psychotic spectrum. Therefore, following studies should recruit a larger sample size to fully cover the variety in SCZ and to be able to identify potential subgroups and biotypes.

### 4.2 Conclusion

The present fMRI study was performed to examine the neural correlates of VM processing in patients with SCZ. Doing so, we used a visual paradigm with three different conditions comprising predictable, random, and arbitrary motion. Across conditions, we identified significantly reduced right TPJ activity in patients with SCZ. This activation deficit is in line with previous studies linking SCZ to impaired VM perception, attentional deficits, and a global sensory processing dysfunction. Due to the latter, the failure to activate the TPJ thus may also reflect an impaired contextual updating process to adjust post-perceptual top-down expectations in patients with SCZ. Across motion conditions patients showed stronger TPJ coupling with higher cognition related brain areas while connectivity with sensory regions was higher in healthy subjects. Given the TPJ’s role as a hub modulating brain network shifting, the aberrant FC may thus reflect a compensatory neural mechanism which co-occurs along with the deficient TPJ activation in SCZ. These findings provide a framework for future research aiming to identify biological markers with a higher potential to inform diagnosis and therapy of the disorder. Therefore, further clinical studies are needed to gain better understanding in the relationship between impaired TPJ activation and aberrant FC in SCZ.

## Data Availability

All data produced in the present study are available upon reasonable request to the authors.

## Acknowledgment

This research project was supported by the START-Program and the Interdisciplinary Center for Clinical Research (IZKF) of the Faculty of Medicine, RWTH Aachen (START 135/14 and ICCR N4-4), the Deutsche Forschungsgemeinschaft (DFG, IRTG-1328) as well as the State of Nordrhein-Westfalen (NRW, Germany) and the European Union through the NRW Ziel2 Program as a part of the European Fund for Regional Development. Furthermore, this work was funded by the Deutsche Forschungsgemeinschaft (DFG,German Research Foundation) – 368482240/GRK2416 (‘RTG 2416 MultiSenses – MultiScales’), by a grant from the Interdisciplinary Centre for Clinical Research within the faculty of Medicine at the RWTH Aachen University (IZKF TN1-8/IA 532008) and the Brain Imaging Facility of the Interdisciplinary Centre for Clinical Research(IZKF) within the faculty of Medicine at the RWTH Aachen University, Germany.

## Author contribution

**Sebastian Scheliga:** Performed data analysis, interpretation, and wrote the manuscript.

**Rosalie Schwank:** Performed data collection and revised the manuscript.

**Ruben Scholle:** Performed data collection and revised the manuscript.

**Ute Habel:** Revised the manuscript and provided critical feedback.

**Thilo Kellermann:** Designed the study, performed data collection, revised the manuscript, and provided critical feedback.

