## Supplementary material for "A neural mechanism underlying predictive visual motion processing in patients with schizophrenia": Captions_Figures

**Figure captions**

**Figure 1.** Illustration of exemplary trajectories of visual motion. Each panel depicts the movement of the black ball roughly during the first 10 seconds of a block. The different panels show exemplary trajectories of the ball during the predictable (A), random (B), and arbitrary (C) motion condition.

**Figure 2.** A) differences in brain activity (yellow blobs with red circles) are displayed between healthy controls (HC) and SCZ patients (SCZ). B) The plot (shows the three VM conditions predictable (Pred), random (Rand), and arbitrary (Arbi). Across conditions, group differences in right TPJ activity are plotted in light grey (Pred), grey (Rand), and dark grey (Arbi).

**Figure 3.** Seed connectivity differences for predictable motion. A) Displayed are brain maps (above) showing significant activation differences in TPJ connectivity between patients and controls. T-values are shown by the colored bar with positive values (from red to yellow) for healthy controls, and negative values (from blue to violet) for patients. B) brain activity is plotted for patients (orange) and healthy controls (blue). Differences in TPJ connectivity are shown for the right lateral occipital cortex (V5), right precuneus, left superior occipital cortex (SOC), and left mPFC.

**Figure 4.** Seed connectivity differences for random motion. A) Displayed are brain maps (above) showing significant activation differences in TPJ connectivity between patients and controls. T-values are shown by the colored bar with positive values (from red to yellow) for healthy controls, and negative values (from blue to violet) for patients. B) brain activity is plotted for patients (orange) and healthy controls (blue). Differences in TPJ connectivity are shown for the mPFC, right SPL, right ITG, left frontal pole, right SFG, and right PcG.

**Figure 5.** Seed connectivity differences for arbitrary motion. A) Displayed are brain maps showing significant activation differences in TPJ connectivity between patients and controls. T-values are shown by the colored bar with negative values (from blue to violet) only for the patients. B) brain activity is plotted for patients (orange) and healthy controls (blue). Differences in TPJ connectivity are shown for the right cerebellum, left SFG, and left PCG.
