## Supplementary material for "A neural mechanism underlying predictive visual motion processing in patients with schizophrenia": Table_S1

| **Patient** | **Medication** | **Doses** |
| --- | --- | --- |
| #1 | Loxapin, Olanzapin, Valproat | NA |
| #2 | Amisulprid | 100mg |
| #3 | Aripiprazol | 30mg |
| #4 | Quetiapin | 1200mg |
| #5 | Clozapin, Citalopram, Aripiprazol | 500mg, 40mg, 10mg |
| #6 | Rivastigmin | NA |
| #7 | Olanzapin | 10mg |
| #8 | - | - |
| #9 | - | - |
| #10 | Amilsulprid | 800mg |
| #11 | - | - |
| #12 | Olanzapin, Aripiprazol | 30mg, 15mg |
| #13 | - | - |
| #14 | - | - |
| #15 | Pipamperon, Doxepin, Aripiprazol | 40mg, 75mg, 20mg |
| #16 | Citalopram | NA |
| #17 | Aripiprazol | NA |
| **Notes**. Four of seventeen patients were free of medication.  **Abbreviations**. NA, not available. | | |

**Table S1.** Medication in the patient group.
