## Supplementary material for "A neural mechanism underlying predictive visual motion processing in patients with schizophrenia": Table_S2

| **Patient** | **Pos.** | **Neg.** | **Gen.** | **Total** |
| --- | --- | --- | --- | --- |
| #1 | 12 | 19 | 24 | 55 |
| #2 | 11 | 7 | 22 | 40 |
| #3 | 21 | 24 | 38 | 83 |
| #4 | 11 | 8 | 19 | 38 |
| #5 | 10 | 21 | 32 | 63 |
| #6 | 9 | 8 | 20 | 37 |
| #7 | 9 | 10 | 17 | 36 |
| #8 | 25 | 14 | 32 | 71 |
| #9 | 9 | 17 | 39 | 65 |
| #10 | 13 | 8 | 26 | 47 |
| #11 | 13 | 14 | 26 | 53 |
| #12 | 8 | 17 | 26 | 51 |
| #13 | 15 | 12 | 26 | 53 |
| #14 | 13 | 15 | 26 | 54 |
| #15 | 14 | 19 | 33 | 66 |
| #16 | 9 | 19 | 28 | 56 |
| #17 | 19 | 8 | 35 | 62 |
| **Abbreviations**. PANSS, positive and negative symptom scale; Pos., positive symptoms subscale; Neg., negative symptoms subscale; Gen., general psychopathology subscale; Total, total PANSS score. | | | | |

**Table S2.** PANSS scores.
